# A phase 3, randomized, controlled trial of Astodrimer 1% Gel for preventing recurrent bacterial vaginosis

**DOI:** 10.1101/2020.08.06.20159475

**Authors:** Jane R. Schwebke, Belvia A. Carter, Arthur S. Waldbaum, Kathy J. Agnew, Jeremy R.A. Paull, Clare F. Price, Alex Castellarnau, Philip McCloud, George R. Kinghorn

**Affiliations:** Division of Infectious Diseases, University of Alabama at Birmingham, Birmingham, AL, USA; Women’s Physician Group, Memphis, TN, USA; Downtown Women’s Health Care, Denver, CO, USA; Department of Obstetrics and Gynecology, University of Washington, Seattle, WA, USA; Starpharma Pty Ltd, Melbourne, VIC, Australia; McCloud Consulting Group, Sydney, NSW, Australia; Royal Hallamshire and Sheffield Teaching Hospitals, Sheffield, United Kingdom

**Author notes:** Author for Correspondence: Jeremy R.A. Paull, PhD, 4-6 Southampton Crescent, Abbotsford, Victoria 3067, Australia. **Conflict of Interest Statement**: JRS is a paid consultant for Starpharma Pty Ltd, Talis One, Toltec, Lupin Pharmaceuticals, and Hologic. ASW received research funding from Gage Development Company. **Financial Disclosure Statement**: This study was funded by Starpharma Pty Ltd. Starpharma Pty Ltd was responsible for the study design, the collection, analysis and interpretation of data, the decision to submit the article for publication, and preparation of the manuscript. The funder provided support in the form of research funding for this study to JRS, BAC and ASW. The funder provided support in the form of salaries for JRAP, CFP and AC, and consulting fees for KJA, PMcC and GRK. **Clinical Trial Registration**: Date of Registration: September 12, 2014; First Patient Enrolled: October 13, 2014; Identification No.: NCT02237950; clinicaltrials.gov. **Paper Presentation**: The 46^th^ Annual Meeting of The Infectious Diseases Society for Obstetrics and Gynecology (IDSOG), Big Sky, Montana, USA, August 8-10, 2019. **Condensation** This phase 3 randomized controlled trial demonstrated efficacy and safety of Astodrimer 1% Gel for the prevention of recurrent bacterial vaginosis.

**Keywords:** Astodrimer Gel, bacterial vaginosis, biofilm, prevention, recurrent bacterial vaginosis, SPL7013, VivaGel

## Abstract

**Objective:** The objective of the study was to confirm the efficacy and safety of Astodrimer 1% Gel to prevent recurrence of bacterial vaginosis.

**Study Design:** 864 women with a diagnosis of bacterial vaginosis and a history of recurrent bacterial vaginosis were enrolled in North America and first received oral metronidazole (500 mg twice daily for 7 days). Women successfully treated with metronidazole were randomly assigned 1:1 to Astodrimer 1% Gel (N=295) or placebo (N=291) at a dose of 5 g vaginally every second day for 16 weeks, and followed for a further 12 weeks off-treatment. The primary endpoint was recurrence of bacterial vaginosis (presence of ≥3 Amsel criteria) at or by Week 16. Secondary endpoints included time to recurrence, and recurrence of subject-reported symptoms. Adverse events were monitored throughout the study.

**Results:** Astodrimer 1% Gel was superior to placebo for the primary and many secondary efficacy measures. At or by Week 16, bacterial vaginosis recurred in 44.2% (130/294) of women receiving astodrimer and 54.3% (158/291) receiving placebo (*P*=.015). Time to recurrence of bacterial vaginosis was significantly longer for women receiving astodrimer compared with placebo (Kaplan-Meier survival curves, *P*=.007). Recurrence of subject-reported symptoms at or by Week 16 was also significantly lower in the astodrimer arm compared with placebo (vaginal odor and/or discharge, 27.9% [75/269] vs 40.6% [108/266], *P*=.002). A significantly lower proportion of patients receiving astodrimer compared with placebo had recurrence of bacterial vaginosis at or by Week 16 by other secondary measures, including individual Amsel criteria (vaginal discharge and clue cells) and Nugent score 7-10. Recurrence of subject-reported vaginal odor and/or discharge was significantly lower in the astodrimer arm compared with placebo up to 8 weeks after cessation of therapy (36.1% [97/269] vs 45.5% [121/266], *P*=.027).

Adverse events were infrequent, and rates were generally similar between placebo and astodrimer groups. Vulvovaginal candidiasis and urinary tract infection occurred more often in women receiving astodrimer.

**Conclusions:** Astodrimer 1% Gel, administered every second day for 16 weeks, was effective and superior to placebo for prevention of recurrent bacterial vaginosis in women with a history of recurrent BV, and was well-tolerated.

## 1 Introduction

Bacterial vaginosis (BV) is the most common vaginal infection and twice as common as vulvovaginal candidiasis.^1^ BV recurrence rates are 43% to 52% within 3 to 6 months of treatment.^2^ BV is a risk factor for serious sequelae, including pre-term birth, and acquisition and transmission of human immunodeficiency virus (HIV) and other sexually transmitted infections (STIs).^3,4^ Recurrent BV has significant psychosocial impacts on women, including severely affecting self-esteem and sex life, and carries a high economic burden.^5,6^

There are no therapies with US regulatory approval for prevention of recurrent BV, and there have been no other adequately powered, well-controlled studies of interventions for recurrent BV. Antibiotic therapies are used off-label over extended periods for reducing recurrence of BV, but are associated with increased risk of side effects^7^ and potential for antibiotic resistance development.^8^ Long-term cure of BV is elusive given the lifestyle factors associated with recurrence.^9^ Therefore, therapies suitable for longer-term use for preventing BV recurrence are urgently required.^10,11^

Astodrimer sodium is a polyanionic dendrimer that blocks attachment of bacteria to cells, preventing formation of bacterial biofilms, which are central to the pathogenesis of BV and not targeted by existing therapies.^12,13,14,15,16^ Astodrimer Gel achieved clinical cure in women with BV following a 7-day treatment course, with 50-74% cured 2-5 days after end of treatment, and was well-tolerated and not systemically absorbed.^17,18^

This phase 3 study assessed Astodrimer 1% Gel for preventing recurrent BV.

## 2 Materials and Methods

### 2.1 Study Design

This was a phase 3, double-blind, multicenter, randomized, placebo-controlled study assessing the efficacy and safety of Astodrimer 1% Gel applied vaginally for 16 weeks compared with placebo (hydroxyethyl cellulose placebo gel) to prevent BV recurrence.

The study complied with the Declaration of Helsinki, was conducted in accordance with Good Clinical Practice, regulatory guidelines, and relevant local legislation, and was approved by an institutional review board on June 20, 2014 (Quorum Review, Inc.). Patient enrolment commenced October 2014 with last follow-up in February 2017.

Patients provided written informed consent and were screened for eligibility at the Screening visit. Eligible patients with a current symptomatic episode of BV and a history of recurrent BV were enrolled in an open-label phase and received oral metronidazole (500mg), twice daily for 7 days. At the second visit (Baseline), 3 to 5 days after completion of metronidazole, women with resolution of BV were randomized 1:1 to either Astodrimer 1% Gel or placebo using a computer-generated randomization list based upon a permutation block procedure.

The active gel and placebo were colorless, clear gels packaged in identical vaginal applicators. Each applicator contained a single dose (5 g) and was individually overwrapped in a sealed pouch. Seventeen applicators (14 doses and 3 spare) were packed in a tamper-evident carton labelled with a unique study medication/patient identification number, allocated using an interactive randomization system. One carton, enough for 4 weeks’ dosing, was dispensed at Baseline, and at Week 4, 8 and 12 study visits. Women self-administered a dose vaginally, every second day for 16 weeks (56 doses total) and attended visits for assessment of BV and adverse events (AEs) every 4 weeks during, and for 12 weeks after end of, treatment. Both care providers and patients were unaware of treatment allocations. Women could withdraw from the study at any time.

Women who had a BV recurrence prior to Week 16 stopped treatment, ended the study and were offered BV therapy as per local practice. A woman was considered to have completed the study if she reached the final follow-up visit (Week 28) recurrence free or had BV recurrence at any time.

### 2.2 Study Population

Women aged 18-45 years with a current diagnosis of BV, defined as presence of ≥3 Amsel criteria (discharge; vaginal fluid pH ≥4.5; ≥20% clue cells; and/or positive 10% potassium hydroxide whiff test),^19^ Nugent score (NS) of 4-10^20^ and self-report of characteristic BV symptoms (abnormal vaginal odor and/or discharge), and a history of recurrent BV defined as ≥2 documented episodes of BV in the past year, were enrolled in the open-label phase.

Women who were pregnant, planning to become pregnant, lactating, or within 3 months of last pregnancy outcome, and women testing positive for urinary tract infection (UTI), or who had signs/symptoms of active genital herpes simplex virus or tested positive for other STIs (*Chlamydia trachomatis, Neisseria gonorrhea* or *Trichomoniasis vaginalis*) at Screening, were excluded.

Women who were asymptomatic and were negative for Amsel criteria for discharge, whiff test and clue cells following the open-label phase, were randomized for the double-blind treatment phase.

Concomitant systemic and vaginal antimicrobial therapies, vaginal antifungals, or any other kind of vaginal products were not permitted during the study.

### 2.3 Outcomes

The primary efficacy endpoint was recurrence of BV at or by Week 16, defined as presence of ≥3 Amsel criteria. Secondary efficacy endpoints included recurrence of subject-reported BV symptoms, individual Amsel criteria, NS 7-10, and ≥3 Amsel criteria and a NS ≥4, at or by Week 16. Time to BV recurrence and BV recurrence at any of the follow-up visits were also secondary efficacy endpoints.

AEs were monitored throughout the study.

### 2.4 Statistical Analyses

Primary and secondary efficacy analyses using logistic regression, with missing BV recurrence data at Week 16 imputed as recurrence, and treatment as the only factor in the model, were performed on the modified intent-to-treat (mITT) population, which comprised all women randomized who administered ≥1 dose of study product. This population was also used to assess safety.

The primary efficacy endpoint was also analyzed for population subgroups, including Nugent category at screening, method of contraception, race, and sexual activity during the study.

Categorical variables were summarized using frequency counts and percentages of patients in each category. Descriptive statistics were calculated for each continuous variable.

Survival curves for time to recurrence were estimated using Kaplan-Meier methodology. A log-rank test was used to test the difference between survival curves of the treatment arms. Hazard of BV recurrence was analyzed within the framework of the Kaplan-Meier survival analysis.

Statistical analyses were performed using SAS (Version 9.2; SAS Institute, Cary, NC).

### 2.5 Sample Size Calculation

Assuming BV recurrence rates of 32% and 45% for Astodrimer Gel and placebo, respectively, a sample size of 308 evaluable participants per treatment arm provided 90% power with a 2-sided test to detect a treatment difference with alpha significance level of 0.05. Therefore, approximately 310 per arm and 620 participants overall were to be randomized into the double-blind treatment phase of the study.

## 3 Results

### 3.1 Disposition and Demographics

A total of 864 women were enrolled in the open-label phase of the study to receive metronidazole; 586/864 (67.8%) eligible women entered the double-blind treatment phase and were randomized to either Astodrimer 1% Gel (N=295) or placebo (N=291) at 67 sites in the US, 4 in Canada, 4 in Mexico and 2 in Puerto Rico. The mITT population included 585 women. Treatment groups were well-balanced with respect to demographic and baseline characteristics (Table 1). The majority of women completed the study and were included in the mITT population (Figure 1).

**Table 1:**
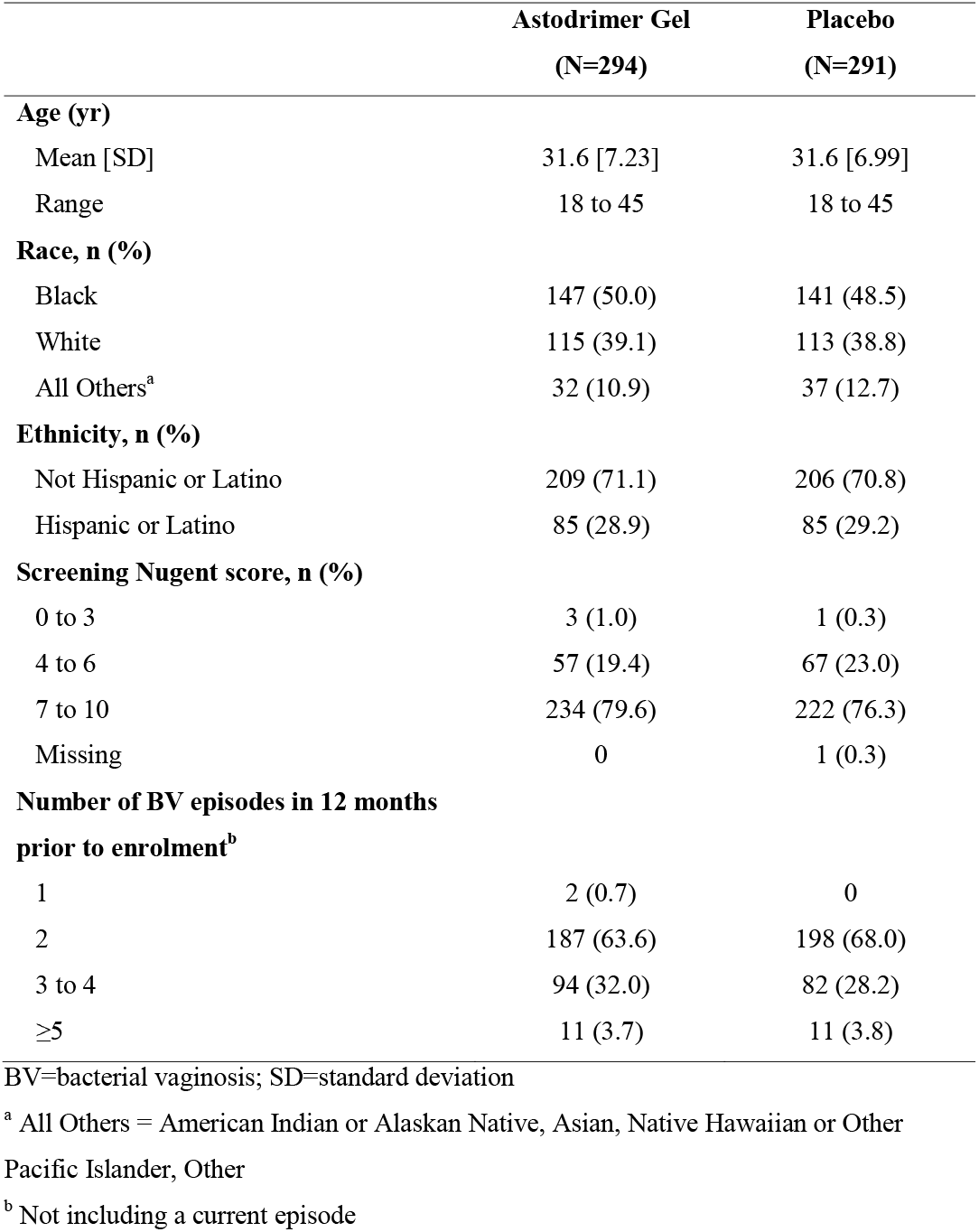
Screening Characteristics (mITT population), by Treatment Group

**Figure 1.**
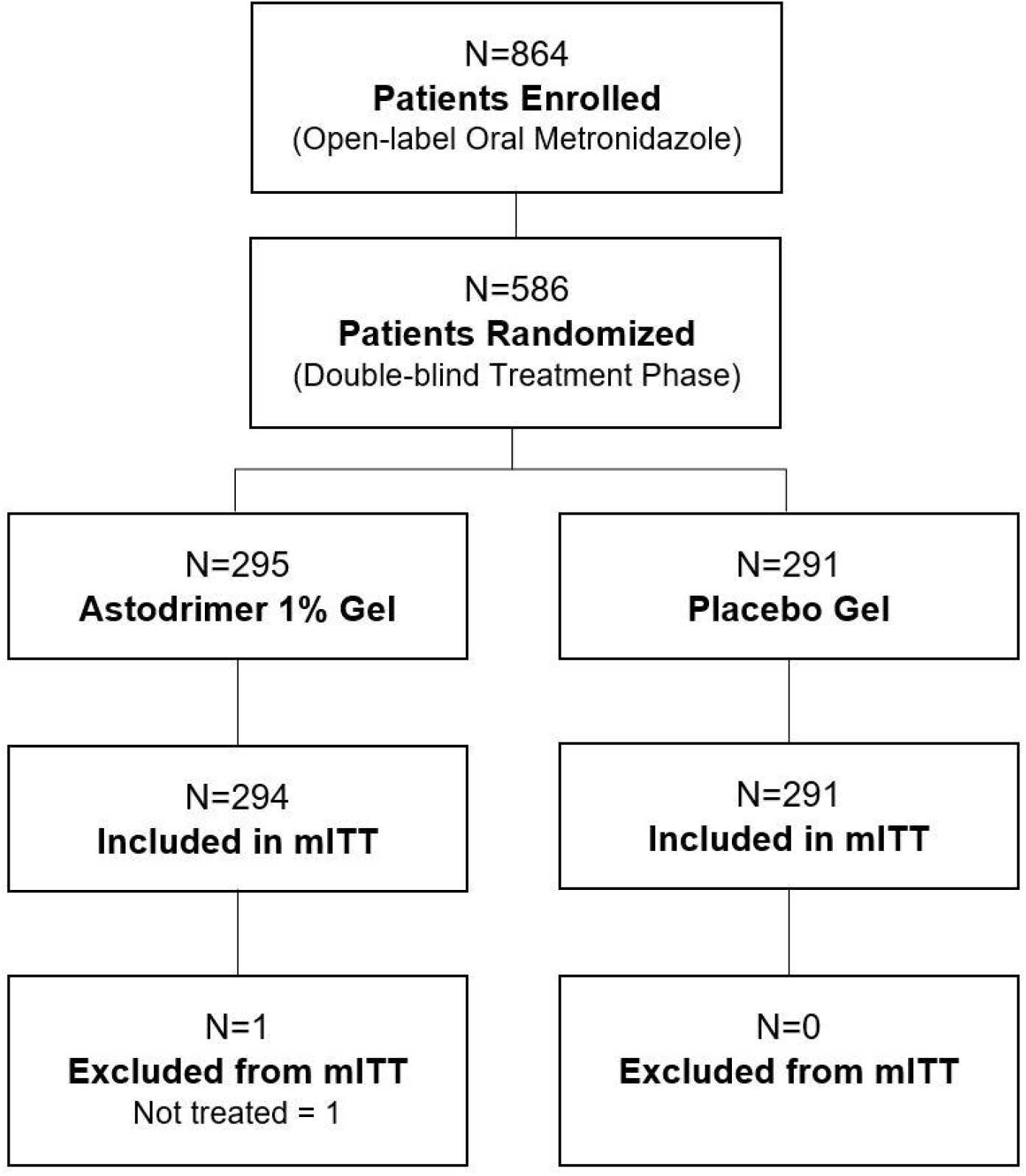
CONSORT diagram

### 3.2 Efficacy

Astodrimer 1% Gel was superior to placebo for the primary endpoint, with 44.2% (130/294) given astodrimer compared to 54.3% (158/291) given placebo experiencing recurrence of BV at or by Week 16; *P*=.015) (Table 2). The recurrence of subject-reported symptoms of BV at or by Week 16 was also significantly lower in the astodrimer arm compared with placebo (Table 2). Kaplan-Meier survival curves for time to recurrence separated after Week 4 and remained so at Week 16; *P*=.007 (Figure 2).

**Table 2:**
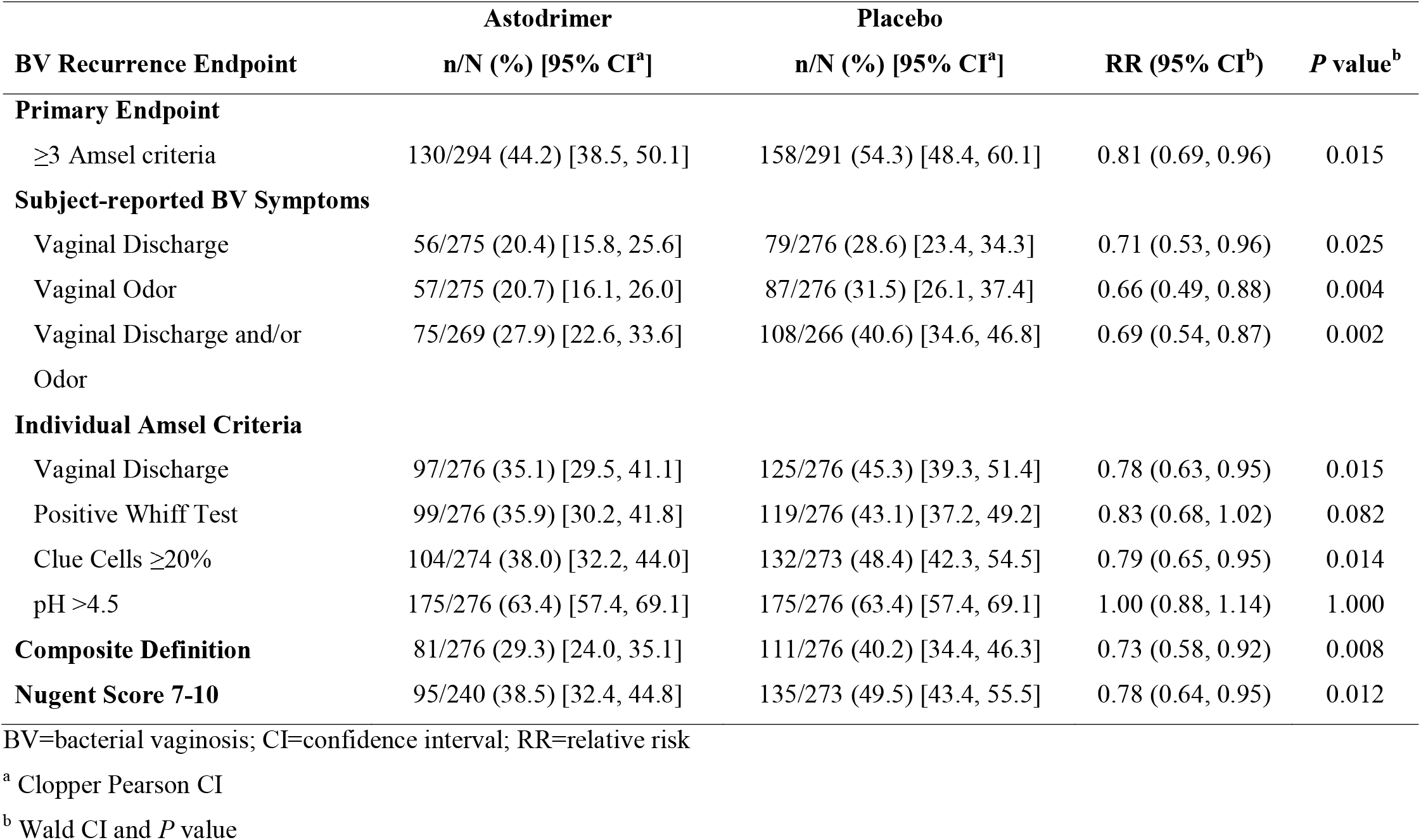
Efficacy Outcomes at or by Week 16 (mITT population), by Treatment Group

**Figure 2.**
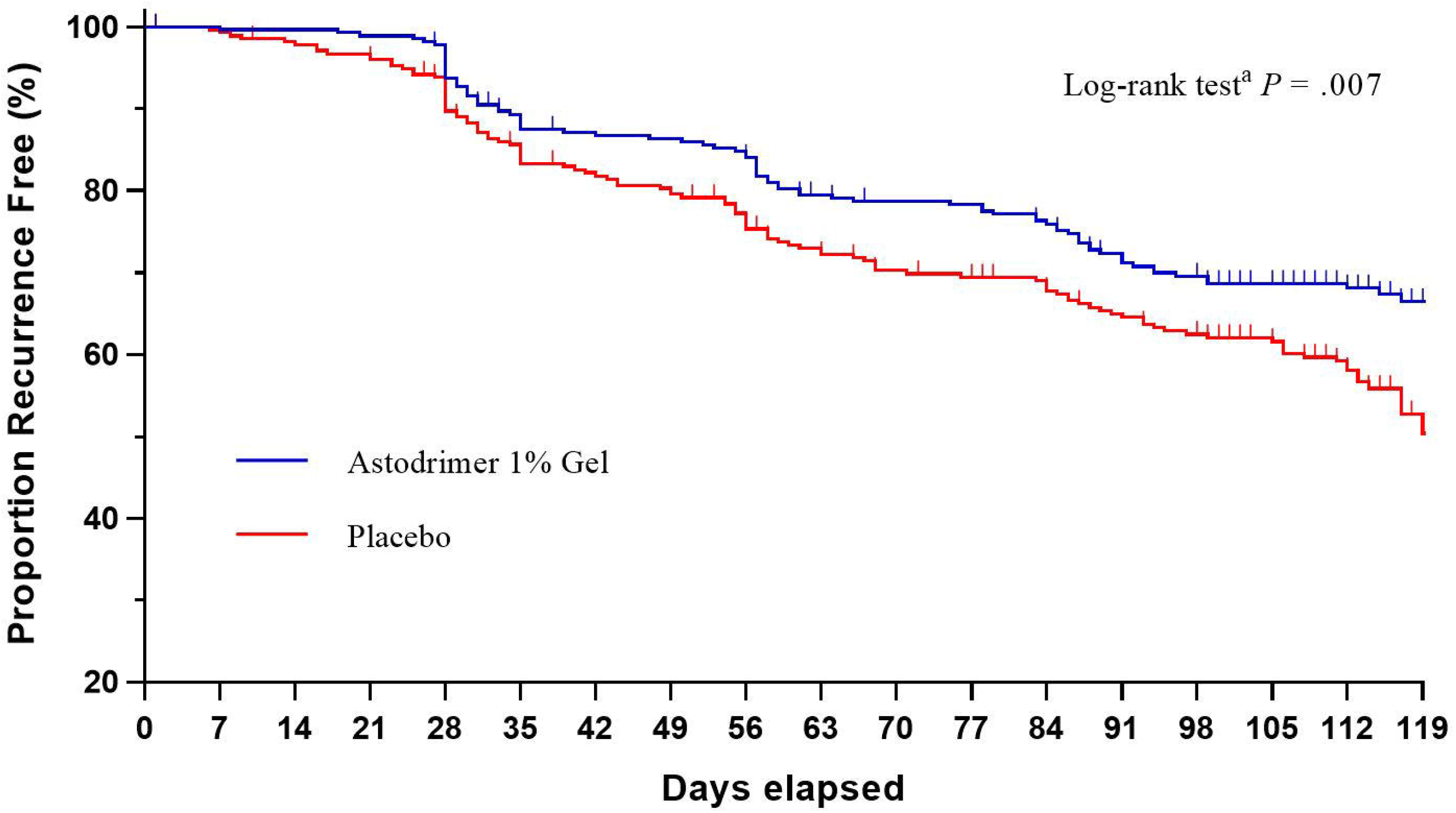
Kaplan-Meier survival curves for time to recurrence of bacterial vaginosis at or by Week 16 (modified intent-to-treat population) ^a^ Log-rank test for the difference between the survival curves of the two treatment groups

Recurrence of all individual Amsel criteria at or by Week 16 was lower in the astodrimer group than in the placebo group, with exception of vaginal fluid pH (Table 2). In addition, a lower proportion of patients receiving astodrimer compared with placebo had BV recurrence at or by Week 16 based on NS 7-10, and the composite of ≥3 Amsel criteria and NS ≥4 (Table 2).

During the 12-week follow-up phase, recurrence of BV (≥3 Amsel criteria) in women given astodrimer was lower than in those given placebo but differences were not statistically significant (Table 3).

**Table 3:**
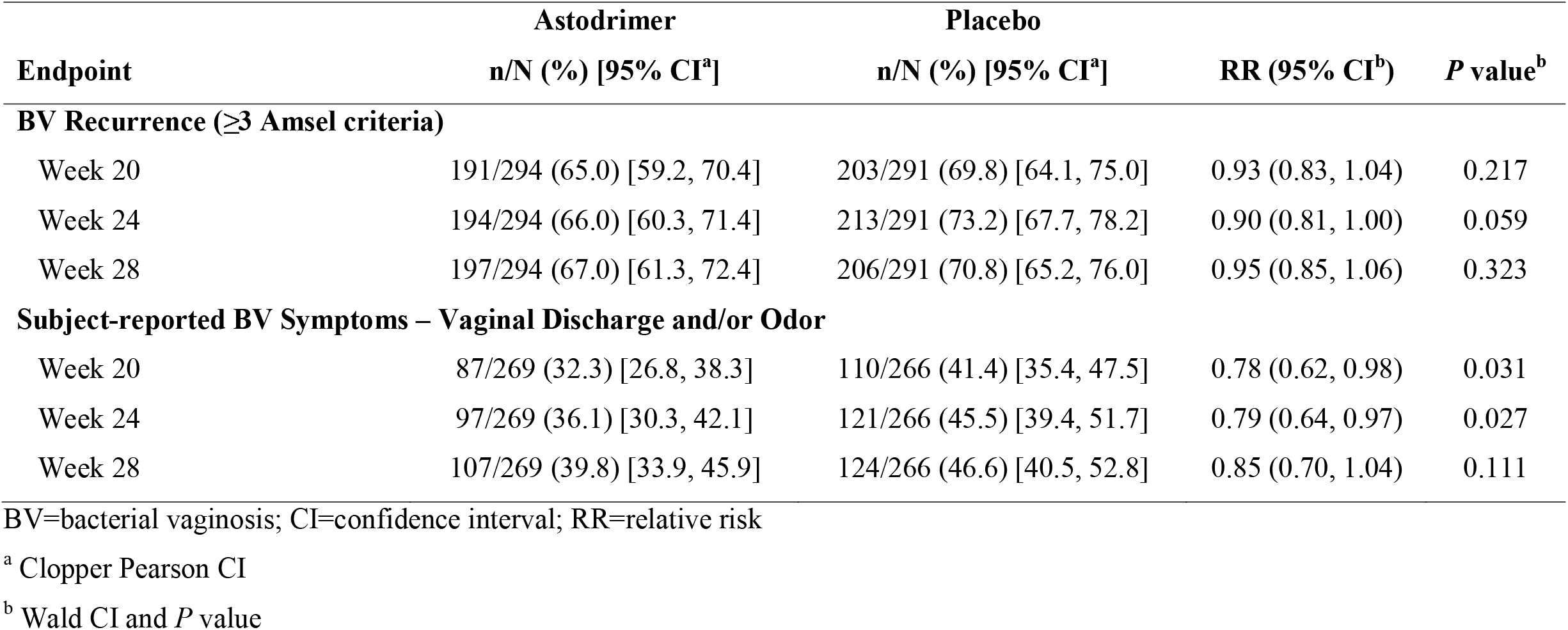
Efficacy Outcomes During Follow-up (mITT population), by Treatment Group

Recurrence of BV symptoms of vaginal odor and/or discharge was statistically significantly lower in the astodrimer arm compared with placebo up to 8 weeks after cessation of therapy (Table 3).

Subgroup recurrence rates were in line with those for the mITT. The Breslow-Day test for homogeneity of odds ratios of astodrimer versus placebo was non-significant for each subgroup factor (*P*>.150, except Age, *P*=.098); data not shown. Lower recurrence at or by Week 16 for women randomized to astodrimer compared to placebo was statistically significant for several subgroup categories, including women with screening NS 7-10 (46.2% [108/234] vs 57.7% [128/222], *P*=.014), black women (53.7% [79/147] vs 68.1% [96/141], *P*=.013), women who had penile-vaginal sexual acts during the treatment period (38.9% [96/247] vs 50.6% [121/239], *P*=.009), and women who used condoms during treatment (33.0% [29/88] vs 49.4% [44/89], *P*=.027).

### 3.3 Safety/tolerability

The overall incidence of AEs was 54.1% (159/294) for Astodrimer Gel and 47.4% (138/291) for placebo (Table 4). AEs potentially treatment-related occurred in 12.6% (37/294) of astodrimer patients and 11.3% (33/291) for placebo.

**Table 4:** Tolerability (mITT population), by Treatment Group

| **Parameter** | **Astodrimer**<br>**N=294**<br>**n (%)** | **Placebo**<br>**N=291**<br>**n (%)** |
| --- | --- | --- |
| Patients with $\geq 1$ AE | 159 (54.1) | 138 (47.4) |
| Patients with $\geq 1$ AE considered by investigator to be potentially related to study treatment | 37 (12.6) | 33 (11.3) |
| Patients with $\geq 1$ severe AE during treatment | 5 (1.7) | 5 (1.7) |
| Patients with $\geq 1$ serious AE | 3 (1.0) | 3 (1.0) |
| Patients who discontinued treatment due to AE | 2 (0.7) | 5 (1.7) |
| Most frequent AEs during treatment (incidence $\geq 2\%$ for astodrimer) | | |
| Vulvovaginal candidiasis | 53 (18.0) | 40 (13.7) |
| Urinary tract infection | 23 (7.8) | 7 (2.4) |
| Headache | 15 (5.1) | 18 (6.2) |
| Abdominal pain (upper, lower, not specified) | 11 (3.7) | 8 (2.7) |
| Vaginal discharge | 9 (3.1) | 4 (1.4) |
| Nasopharyngitis | 8 (2.7) | 13 (4.5) |
| Upper respiratory tract infection | 8 (2.7) | 6 (2.1) |
| Vulvovaginal pruritus | 7 (2.4) | 10 (3.4) |

AE=adverse event

Most AEs were mild or moderate in intensity, and self-limiting. During treatment, 1.7% (5) women in each group reported severe AEs; none were considered treatment related. In the astodrimer group, 1 participant discontinued treatment due to menorrhagia and 1 participant due to vulvovaginal candidiasis, which was considered possibly related to treatment. For placebo, 1 participant discontinued due to each of vulvovaginal candidiasis, type 2 diabetes mellitus, headache and abdominal pain, and 1 participant discontinued after experiencing vaginal inflammation, vulvovaginal burning sensation, vulvovaginal pruritus and BV considered possibly treatment-related.

Serious AEs were reported for 3/294 (1.0%) women in the astodrimer group and 3/291 (1.0%) in placebo, and none was considered to be potentially treatment related.

Vulvovaginal candidiasis was reported in 18.0% (53/294) and 13.7% (40/291) women in the astodrimer and placebo groups, respectively, during treatment and 20.1% [59/294] vs 17.2% [50/291] for the overall study period. Vulvovaginal candidiasis considered potentially treatment-related was reported in 6.8% (20/294) and 4.8% (14/291) of women using astodrimer or placebo, respectively, during treatment. During follow-up, vulvovaginal candidiasis rates were 4.1% (12/294) for astodrimer and 5.8% (17/291) for placebo. During treatment, UTI rates were 7.8% (23/294) for astodrimer and 2.4% (7/291) for placebo.

## 4 Discussion

Astodrimer 1% Gel, administered every second day for 16 weeks, was effective and superior to placebo for the prevention of BV recurrence in women with a history of recurrent BV. The primary efficacy finding was supported by multiple secondary endpoints, including significantly longer time to recurrence and lower recurrence of symptoms, which was significant up to 8 weeks after end of treatment.

Astodrimer 1% Gel was well-tolerated, with the incidence of AEs generally similar between the astodrimer and placebo arms. Rates of candidiasis were generally low.

The current study of Astodrimer 1% Gel represents the largest and first adequately powered, randomized, placebo-controlled study of a therapy for preventing recurrent BV.

Some approved antibiotics or investigational therapies have been shown to reduce recurrence of BV in limited clinical studies that have been generally non-randomized, not placebo-controlled, and/or not adequately powered.^7,21,22,23^ Nevertheless, off-label regimens of products not approved for prevention, including topical metronidazole and topical acid boric over periods of 4-6 months, together with fluconazole to prevent likely secondary candidiasis, are recommended for reducing recurrent BV.^24^

The proportion of women with known BV recurrence (i.e., missing data not imputed) for Astodrimer 1% Gel (34.9% [88/252] vs 46.6% [116/249] for placebo) was comparable with that seen in a similarly designed study of topically applied metronidazole gel given for 16 weeks (25.5%).^7^

The difference in recurrence rates between astodrimer and placebo narrowed progressively after end of treatment, but women in the astodrimer group recurred later than placebo, and therefore had more recurrence free days. In addition, recurrence of symptoms of BV was statistically significantly lower for astodrimer compared with placebo up to Week 24, 8 weeks after end of therapy, indicating a clinically meaningful residual benefit.

Treatment with astodrimer helped maintain normal vaginal flora, with lower BV recurrence rates as determined by Nugent score and the combination of Amsel criteria and Nugent score.

The proportion of women with vulvovaginal candidiasis receiving astodrimer (18% during treatment, 20.1% overall) was similar to placebo and less than half that reported during a study of 16 weeks’ treatment with topical metronidazole 0.75% gel (43.1% vs 20.5% in placebo).^7^

The slightly higher proportion of women with UTI for astodrimer compared with placebo could be potentially explained by the longer symptom-free period associated with use of astodrimer allowing a resumption of a more normal frequency of intercourse, consistent with slightly higher sexual functioning scores observed in this study. In any case, the incidence of uncomplicated UTI in young, sexually active women in the US is reported to be approximately 0.5 episodes per person per year.^25^ The incidence in women using astodrimer in this study was 0.37 episodes per person per year.

There were no other notable differences in AEs reported for astodrimer and placebo.

Astodrimer is a novel dendrimer administered vaginally and is not systemically absorbed. Data show that it inhibits formation of and disrupts biofilms due to its ability to block bacterial adhesion. Given this profile, astodrimer avoids issues typically associated with conventional antibiotics, such as systemic side effects and antibiotic resistance, and is suitable as an effective and safe alternative for the long-term management of recurrent BV, addressing an unmet medical need.

Subgroup analyses showed statistically significant differences in clinical response between astodrimer and placebo in population groups with recognized risk factors for BV, such as black women, those engaging in penile-vaginal sexual acts during the treatment period, and a high screening NS of 7-10, suggesting benefit of preventive treatment with astodrimer in these high-risk groups. Subgroup analyses were exploratory in nature and results should be interpreted with caution.

The findings of this Phase 3 study support the clinical utility of Astodrimer 1% Gel as a novel treatment for prevention of BV recurrence in women suffering from recurrent BV. The product has regulatory approval in Europe, Australia and a number of countries in Asia, and additional safety information will be derived from routine post-market surveillance activities.

The study is the largest randomized, double-blind, placebo-controlled study of a therapy to prevent recurrent BV and was adequately powered to detect a difference in rates of BV recurrence between astodrimer- and placebo-treated women.

The results of reduced recurrence of BV at Week 16 are consistent with the ability of Astodrimer 1% Gel to achieve clinical cure of BV at the end of a 7-day treatment period, as demonstrated in phase 2 and 3 clinical studies.^17,18^

## 5 Conclusions

The study supports a role for Astodrimer 1% Gel as an effective long-term therapy to prevent recurrence of BV, with a novel mechanism of action related to blocking of biofilms. The product is not systemically absorbed, and offers patients and clinicians a unique treatment option that avoids potential issues associated with existing antibiotics.

## Data Availability

The data that support the findings of this study are available from the corresponding author (JRAP) upon reasonable request.

## Acknowledgments

The authors would like to thank all the patients who were involved in the study and the Principal Investigators for involvement with the clinical trial. Editorial and medical writing assistance was provided by Angela Hart, MSc, of Quanticate UK Ltd, supported by Starpharma Pty Ltd, the manufacturer of Astodrimer Gel. The authors were fully responsible for the content, editorial decisions, and opinions expressed in the current article. The authors did not receive an honorarium related to the development of this manuscript.

## References

1 Barbone F, Austin H, Louv WC, Alexander WJ. A follow-up study of methods of contraception, sexual activity, and rates of trichomoniasis, candidiasis, and bacterial vaginosis. Am J Obstet Gynecol 1990;163(2):510–4.

2 Bradshaw CS, Morton AN, Hocking J, et al. High recurrence rates of bacterial vaginosis over the course of 12 months after oral metronidazole therapy and factors associated with recurrence. J Infect Dis 2006;193:1478–86.

3 Hillier SL, Nugent RP, Eschenbach DA, et al. Association between bacterial vaginosis and preterm delivery of a low-birth-weight infant. The Vaginal Infections and Prematurity Study Group. N Engl J Med 1995;333:1737–42.

4 Schwebke JR. New concepts in the etiology of bacterial vaginosis. Current Infectious Disease Reports 2009;11(2):143–7.

5 Bilardi JE, Walker S, Temple-Smith M, et al. The burden of bacterial vaginosis: Women’s experience of the physical, emotional, sexual and social impact of living with recurrent bacterial vaginosis. PLoS ONE 2013;8(9):e74378.

6 Peebles K, Velloza J, Balkus JE, McClelland RS, Barnabas RV. High global burden and costs of bacterial vaginosis: A systematic review and meta-analysis. Sex Transm Dis 2019;46(5):304–11.

7 Sobel JD, Ferris D, Schwebke J, et al. Suppressive antibacterial therapy with 0.75% metronidazole vaginal gel to prevent recurrent bacterial vaginosis. Am J Obstet Gynecol 2006. 194:1283–9.

8 Beigi RH, Austin MN, Meyn LA, Krohn MA, Hillier SL. Antimicrobial resistance associated with the treatment of bacterial vaginosis. Am J Obstet Gynecol 2004;191:1124–9.

9 Bradshaw CS, Vodstrcil LA, Hocking JS, et al. Recurrence of bacterial vaginosis is significantly associated with posttreatment sexual activities and hormonal contraceptive use. Clin Infect Dis 2013;56(6):777–86.

10 Bradshaw CS, Brotman RM. Making inroads into improving treatment of bacterial vaginosis – striving for long-term cure. BMC Infect Dis 2015;15:292–303.

11 Bradshaw CS, Sobel JD. Current treatment of bacterial vaginosis - Limitations and need for innovation. J Infect Dis 2016;214(Suppl 1):S14–20.

12 Swidsinski A, Mendling W, Loening-Baucke V, et al. Adherent biofilms in bacterial vaginosis. Obstet Gynecol 2005;106(5 Pt 1):1013–23.

13 Muzny CA, Schwebke JR. Biofilms: An underappreciated mechanism of treatment failure and recurrence in vaginal infections. Clin Infect Dis 2015;61(4):601–6.

14 Machado D, Castro J, Palmeira-de-Oliveira A, Martinez-de-Oliveira J, Cerca N. Bacterial vaginosis biofilms: Challenges to current therapies and emerging solutions. Front Microbiol 2016;6:1528.

15 McGowan I, Gomez K, Bruder K, et al. Phase 1 randomized trial of the vaginal safety and acceptability of SPL7013 Gel (VivaGel) in sexually active young women (MTN-004). AIDS 2011;25(8):1057–64.

16 O’Loughlin J, Millwood IY, McDonald HM, Price CF, Kaldor JM, Paull JRA. Safety, tolerability, and pharmacokinetics of SPL7013 Gel (VivaGel): A dose-ranging, phase I study. Sex Transm Dis 2010;37(2):100–4.

17 Waldbaum AS, Schwebke JR, Paull JRA, et al. A phase 2, double-blind, multicenter, randomized, placebo-controlled, dose-ranging study of the efficacy and safety of Astodrimer Gel for the treatment of bacterial vaginosis. PLoS ONE 2020;15(5):e0232394.

18 Chavoustie SE, Carter BA, Waldbaum AS, et al. Two phase 3, double-blind, placebo-controlled studies of the efficacy and safety of Astodrimer 1% Gel for the treatment of bacterial vaginosis. Eur J Obstet Gynecol Reprod Biol 2020;245:13–8.

19 Amsel R, Totten PA, Spiegel CA, Chen KCS, Eschenbach D, Holmes KK. Nonspecific vaginitis; Diagnostic criteria and microbial and epidemiologic associations. Am J Med 1983;74:14–22.

20 Nugent RP, Krohn MA, Hillier SL. Reliability of diagnosing bacterial vaginosis is improved by a standardized method of gram stain interpretation. J Clin Microbiol 1991;29:297–301.

21 Reichman O, Akins R, Sobel JD. Boric acid addition to suppressive antimicrobial therapy for recurrent bacterial vaginosis. Sex Transm Dis 2009;36(11):732–4.

22 McClelland RS, Balkus JE, Lee J, et al. Randomized trial of periodic presumptive treatment with high-dose intravaginal metronidazole and miconazole to prevent vaginal infections in HIV-negative women. J Infect Dis 2015;211(12):1875–82.

23 Cohen CR, Wierzbicki MR, French AL, et al. Randomized trial of lactin-V to prevent recurrence of bacterial vaginosis. N Engl J Med 2020;382:1906–15.

24 Workowski KA, Bolan GA. Centers for Disease Control and Prevention (CDC). Sexually transmitted diseases treatment guidelines. MMWR-Morbid Mortal W 2015;64(RR-03):1–137.

25 Medina M, Castillo-Pino E. An introduction to the epidemiology and burden of urinary tract infections. Ther Adv Urol 2019;11:3–7.

